# Phenomic Network Analysis for Depression Reveals Comorbidity Architecture, Genomic Relationships, and Pleiotropic Variants

**DOI:** 10.1101/2022.05.13.22275045

**Authors:** Zhiyu Yang, Pritesh Jain, Petros Drineas, Peristera Paschou

## Abstract

Depression is one of the most prevalent psychiatric disorders and is one of the leading causes of health ailment worldwide. It is known to be highly heritable and is frequently comorbid with other mental and physical traits. This observation motivated us to look deeper into the genetic and phenotypic connections between depression and other traits in order to identify correlations as well as potentially causal connections between them. In this study, we analyzed data from the UK biobank to systematically evaluate relationships between depression and other heritable traits both from a phenotypic and a genetic aspect. We compressed a total of 6,300 ICD codes into 412 heritable phecodes and we constructed a comorbidity network connecting depression and other disorders on over 300,000 participants of European ancestry. Additionally, we investigated the genetic correlation for each (phenotypic) connection in the resulting network. We also looked into potentially causal relationships using mendelian randomization for all pairs of significantly correlated disorders and uncovered horizontal pleiotropic genetic variants and genes contributing to disease etiologies. We found gastro-oesophageal reflux disease (GORD), body mass index, and osteoarthritis to be direct causes for depression, with GORD lying at the center of the causal network. Genes broadly expressed in various tissues, such as *NEGR1, TCF4*, and *BTN2A1* underlie the pathways that lead not only to depression but also to other related disorders. Our work highlights the broad connections between depression and diverse traits, indicating a complex etiology and possible existence of subtypes for depression. Our findings highlight the value of cross-trait analysis in order to better understand the neurobiology of complex psychiatric disease.

## 1 Introduction

As one of the most prevalent psychiatric disorders, depression is a major burden on global health. Currently, it is affecting approximately 10% of the adult population in the United States (22) and shows an increasing trend in its incidence rate across the world (57; 69). According to the fifth edition of Diagnostic and Statistical Manual of Mental Disorders (DSM-V), a diagnosis of a major depressive episode requires at least one main symptom of depressed mood or *anhedonia* (defined as the loss of interest or pleasure), along with some secondary symptoms, including disturbance in appetite, sleep, or ability to engage in physical activity and cognition (1). However, despite the existence of precise diagnostic criteria, a high level of heterogeneity has been observed among depressive patients; this is common in many psychiatric traits that are nevertheless managed as dichotomous.

Individuals diagnosed with depression often also present with other mental and physical illness. On the psychiatric side, traits commonly co-occuring with depression include anxiety (97; 48), substance abuse (16), and post-traumatic stress disorder (PTSD) (28; 46). Meanwhile, depression is also reported to be highly comorbid with certain clinically distinct phenotypes, such as obesity (19; 4), cardiovascular diseases (41; 75), and even immune disorders including multiple sclerosis (MS) (30; 60) and inflammatory bowel disease (IBD) (62; 2). Given the wide range of phenotypes coexisting with depression, it is important to quantify and evaluate these comorbidities in depression patients systematically. However, perhaps due to lack of resources, such studies for depression are very limited. Most existing studies focusing on depression’s comorbidity targeted only a small group of homogeneous diseases rather than a broader spectrum (36; 39; 99; 37; 40). Few studies aimed at recovering a more general comorbidity network of depression (84; 96; 63) but stopped at an epidemiological level, without further exploring underlying biological mechanisms. On the other hand, existing phenome-wide association studies (PheWAS) attempted to evaluate the association between various phenotypes and the genetic risk of depression (72). However, using an individual’s genetic risk score as a proxy may result in limited clinical interpretability.

The emergence of large comprehensive biobanks, as well as the abundance of publicly available genome-wide association studies (GWAS), allows us to systematically investigate depression and its comorbidities from both a phenotypic and a genetic perspective. Depression as well as most of the aforementioned mentioned co-occurring disorders have strong genetic foundations (89; 101; 64; 88; 38), with depression itself showing a heritability of approximately 30%-40% in family-based studies (32; 65; 91) and a SNP heritability of approximately 10% (55; 43). In this study, we investigate both the phenotypic and genetic architecture of depression and its comorbid disorders, leveraging the UK biobank datasets and largest available genomewide association studies (GWAS) to construct phenotypic and genetic networks. Our results shed light on the epidemiological and phenotypic relationships between depression and other disorders and help uncover biological insights that may underlie the clinically observed comorbidities.

## 2 Methods

### 2.1 Data

The UK Biobank dataset includes phenotypic and genetic data from approximately 500,000 individuals and was used to compute phenotypic correlations. As a quality control step, we removed samples with non-European British ancestry, based on their self-reported ancestry information. Next, we removed individuals with greater than third degree relatedness based on the kinship coefficient. Finally, we ran Principal Component Analysis (PCA) using TeraPCA (11) to remove individuals who do not overlap with European samples in the 1000 genomes dataset (mean principal component value on the top six principal components plus/minus three standard deviations). We further removed individuals with genotype missingness less than 0.98, thus retaining a dataset of 331,256 participants.

For the phenotypic data, we included 6,300 ICD-10 diagnoses (data field 41270) with a non-zero number of patients in our analyses. To reduce the dimensionality and increase the interpretability of our analyses, we further mapped the ICD-10 codes on PheCodes. Out of all 6,300 ICD10 diagnoses, 4,807 were mapped to at least one valid PheCode, for a total of 505 PheCodes and 1,434 child PheCodes. We removed PheCodes from categories that are dominated by non-genetic causes (infectious diseases, injuries, poisonings, pregnancy complications, etc.). This resulted in a total of 4,004 ICD-10 diagnoses mapped onto 411 parent PheCodes. We decided to separate depression from its parental PheCode 296 (mood disorders), which included bipolar and other mood disorders. We ended up with 412 phenotypic end points, including 410 parental PheCodes, plus one for depression and one for other mood disorders. (See Table S1 for the detailed list.)

### 2.2 Phenotypic network of depression and comorbid disorders

We computed phenotypic correlations between all pairs of PheCodes, a total of 412×411/2 = 84, 666 pairs. Out of all these possible combinations, we observed 77,502 non-zero phenotypic correlations, where phenotypic correlation is defined as cosine of the angle formed between pairs of phenotype vectors. We focused our analyses on highly comorbid pairs of disorders that show phenotypic correlations in the top 1% quantile, i.e., we focused on the top 775 pairs. We extracted depression and its comorbid disorders from those top 775 pairs and we formed a phenotypic correlation network. For each trait that was deemed comorbid with depression, we used a hypergeometric test to analyze its enrichment in cases of depression patients.

### 2.3 Genetic network across depression and its comorbidities

To study the genetic relationship between depression and its comorbid disorders, we used linkage disequilibrium (LD) score regression (as implemented in LDSC (14)) to evaluate genetic correlations between all highly comorbid pairs in our network. For this analysis, we obtained publicly available summary statistics from GWAS of identical or similar traits (see Table 1). In order to retain independence between our phenotypic and genetic networks, we selected GWAS summary statistics that did not involve samples from the UK biobank. However, this was not possible for certain disorders (see Table 1). For all genetic correlation analyses, only SNPs that were matched with the HapMap3 SNP list were used. As the GWAS summary statistics are dominated by European samples, we used LD scores estimated from the European subgroup of the 1000 Genomes phase 3 project as both the independent variable and the regression weights. The significance threshold for this analysis was corrected by dividing by the number of tested pairs, which corresponds to the number of highly comorbid pairs within the phenotypic correlation network.

**Table 1:** Sources for GWAS summary statistics. Summary statistics are dominated by samples of European ancestry, with two exceptions (hypertesion and sodium/potassium levels).

| Trait | Reference | Sample Size | comment |
| --- | --- | --- | --- |
| asthma | (26) | 23,948 cases, 118,538 controls |  |
| body mass index | (42) | 100,418 samples |  |
| gastro-oesophageal reflux | (6) | 80,265 cases, 305,011 controls |  |
| hypertension | (105) | 27,123 cases, 22,018 controls | diverse population |
| osteoarthritis | (108) | 30,727 cases and 297,191 controls |  |
| depression | (106) | 59,851 cases and 113,154 controls |  |
| hernia | (102) | 14,278 cases, 286,513 controls |  |
| anxiety | (80) | 7,016 cases, 14,745 controls |  |
| low-density lipoprotein | (103) | 94,595 samples |  |
| high-density lipoprotein |  | 94,595 samples |  |
| sodium levels | (49) | 6,018 samples | Qatar population |
| potassium levels |  | 6,017 samples |  |
| inflammatory bowel disease | (23) | 25,042 cases, 34,915 controls |  |
| suicide attempt | (102) | 125,844 samples |  |
| chronic gastritis | (102) | 15,492 cases, 229,398 controls |  |

### 2.4 Phenome-wide causal inference

For all disorder pairs where a significant genetic correlation was observed, we further investigated their potential causative relationship using bidirectional, generalized, summary-statistic-based Mendelian randomization (GSMR) (111). For each analysis, we used SNPs that were associated with the exposure trait (*p*-value less than 5 × 10−6) as instrument variables; we required a minimum of five independent SNPs (*r*^2^ < 0.05) in this analysis. We excluded pleiotropic SNPs identified through the heterogeneity in the dependent instrument (HEIDI outlier approach, SNPs with *p*_*HEIDI*_ < 0.01) to guarantee that all instrument SNPs affect the outcome only via exposure. We used Bonferroni correction, correcting for the number of significant genetic correlations (multiplied by two for bidirectional tests) to determine the significance threshold of this analysis.

### 2.5 Evaluation of horizontal pleiotropy

We further looked into genetic variants that show horizontal pleiotropy effects between depression and disorders that appeared to be causally related to depression from the Mendelian randomization analysis. The analysis was carried out using Iterative Mendelian Randomization and Pleiotropy (IMRP) (110). Using causal effect sizes and standard errors estimated from the afore-mentioned Mendelian randomization analyses, we ran SNP-based pleiotropic test for depression and each causal exposure respectively. Similar to mendelian randomization, the test was carried out in a bi-directional manner. This is equivalent to running a GWAS for two traits: *β*_*pleio*1_ = *β*_*depression*_–*β*_*exp*−*dep*_ × *β*_*exposure*_ and *β*_*pleio*2_ = *β*_*exposure*_–*β*_*dep*−*exp*_ × *β*_*depression*_, where *β*_*depression*_ and *β*_*exposure*_ stand, respectively, for GWAS effect sizes for depression and the causal exposure; and *β*_*exp*−*dep*_ and *β*_*dep*−*exp*_ are the estimated causal effect sizes from the exposure trait to depression and from depression to the exposure trait, respectively. To identify pleiotropic SNPs between depression and the exposure trait, we first selected SNPs that were genome-wide significant (*p* < 5 × 10^−8^) in either direction. For a SNP to be considered pleiotropic, it has to satisfy two conditions: first, it must be genome-wise significant in one direction and, second, it must have a *p*-value below the Bonferroni-corrected threshold in the other direction. In more detail, suppose that we identified a total of *n* SNPs being genome-wide significant in either direction. Then, the pleiotropic SNPs will be the ones that have *p* < 5 × 10^−8^ in one direction and *p* < 0.05*/n* in the other direction.

On top of the SNP-based pleiotropic test, we also seek to interpret the results at a gene level. Therefore, to analyze depression-exposure pairs, we first carried out gene-based analyses using results from SNP-based pleiotropic tests for both directions using MAGMA (24). We subsequently performed gene-based pleiotropic screening on the MAGMA results. In this case, significant pleiotropic genes were defined as ones that are genome-wise significant (*p* < 0.05 divided by the total number of genes tested for the trait) in one direction *and* show *p*-value lower than a Bonferroni-corrected significance threshold (< 0.05 divided by the total number of genes satisfying the first condition) in the other direction.

## 3 Results

### 3.1 Phenotypic network of depression and its comorbidities

Among the 331,256 unrelated European samples that we retained from the UK biobank, 18,588 have been diagnosed with depression. Out of the 412 endpoint traits that we defined, we identified 12 frequent comorbidities with depression among the top 1% quantile of all pairwise phenotypic correlations. The most frequent one is hypertension (parent PheCode 401), with nearly half of all depression patients having been diagnosed with hypertension (*N*_*Hypertension*|*Depression*_ = 9, 196). The next trait most frequently comorbid to depression, was osteoarthrosis (parent PheCode 740), which was observed in more than one-third of all depression patients (*N*_*Osteoarthrosis*|*Depression*_ = 6, 902). Interestingly, out of the top 12 comorbidities for depression in the UK Biobank, only two are psychiatric or behavioral traits: anxiety disorders (parent PheCode 300, *N*_*Anxiety*|*Depression*_ = 5, 666) and suicidal ideation or attempt (parent PheCode 297, *N*_*Suicide*|*Depression*_ = 1, 154). Even though the number of comorbid cases with depression for either of these two psychiatric traits is not as high as hypertension or osteoarthrosis, one has to consider anxiety disorders and suicidal ideation are not as prevalent in the UK biobank. Anxiety disorders and suicidal ideation are highly enriched within depression patients (*p*_*Anxiety*_ = 5.84 × 10^−3711^ and *p*_*Suicide*_ = 2.56 × 10^−1120^). The remaining eight traits span conditions that affect various organs, including gut, lungs, and processes of the metabolic system (see Table 2). Moreover, just as expected, all these comorbidities of depression are also tightly connected to each other (see Figure 1 and Table S2).

**Table 2:** Enrichment of comorbidities in depression patients. For all traits phenotypically correlated with depression (Figure 1), we tested the enrichment of cases in depression patients using a hyper-geometric test. Enrichment *P* denotes the hyper-geometric test *p*-value; total *N*_*sample*_ denotes the total number of cases of the trait in the UK biobank samples; *N*_*sample*_ in depression denotes the number of the cases having the respective trait *and* depression.

| Trait | total $N_{sample}$ | $N_{sample}$ in depression | Enrichment $P$ |
| --- | --- | --- | --- |
| All Sample | 331,256 | 18,588 | - |
| Depresssion | 18,588 | 18,588 | - |
| Disorder of lipid metabolism | 46,738 | 5,394 | $1.84 \times 10^{-644}$ |
| Osteoarthrosis | 59,572 | 6,902 | $1.99 \times 10^{-887}$ |
| Diseases of esophagus | 47,027 | 5,664 | $2.09 \times 10^{-764}$ |
| Suicidal ideation or attempt | 1,495 | 1,154 | $2.56 \times 10^{-1120}$ |
| Asthma | 29,737 | 3,892 | $3.10 \times 10^{-578}$ |
| Abdominal hernia | 54,605 | 5,297 | $3.34 \times 10^{-391}$ |
| Functional digestive disorders (IBD) | 31,818 | 4,342 | $4.09 \times 10^{-711}$ |
| Gastritis and duodenitis (ChronicGastritis) | 34,101 | 4,188 | $4.16 \times 10^{-550}$ |
| Fluid and electrolyte disorders | 16,418 | 2,879 | $4.25 \times 10^{-685}$ |
| Overweight obesity and other hyperalimentation | 21,266 | 3,529 | $4.26 \times 10^{-787}$ |
| Anxiety disorders | 13,186 | 5,666 | $5.84 \times 10^{-3711}$ |
| Hypertension | 94,738 | 9,196 | $7.48 \times 10^{-833}$ |

**Figure 1:**
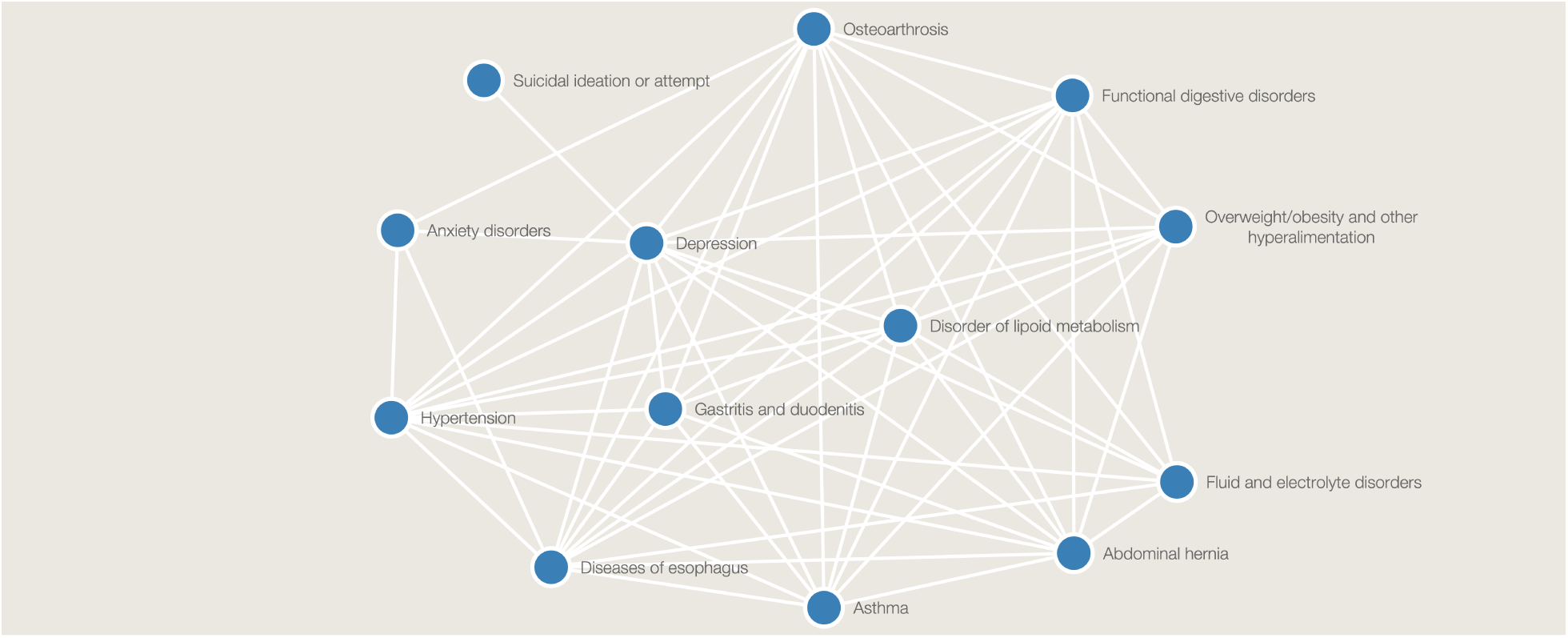
Phenotypic disease network of depression. The disease network of depression was obtained from the fully connected phenotypic network using phenoytpic correlations as edge weights, by reducing the dense graph to a sparse network by keeping only the top 1% of the connections and extracting depression and its immediate neighbour nodes.

### 3.2 Genetic network of depression and its comorbidities

We observed a total of 56 connections in the phenotypic network of depression and its 12 comorbid traits discussed above. We further proceeded to examine the potential genetic relationships between disorder pairs. Focusing on the 13 traits in the phenotypic network, we were able to find publicly available GWAS summary statistics for all of them (see Table 1). For lipid metabolism disorders (parent PheCode 277), we used two sets of summary statistics, one corresponding to low-density lipoprotein levels and one corresponding to high-density lipoprotein levels. Similarly, for fluid and electrolyte disorders (parent PheCode 276), we also used two sets of summary statistics as proxies, one corresponding to sodium level measurements and one corresponding to potassium level measurements. These resulted in a total of 76 independent tests for genetic correlations and a Bonferroni-corrected significance threshold less than 6.58 × 10^−4^. Using this threshold, we identified 18 genetically correlated pairs of disorders, resulting to a fully connected genetic network (see Table 3 and Figure 2). We found that even though at the phenotypic level all 12 traits are highly comorbid with depression, only six out of the 12 actually share a genetic basis with depression (see Table 3). More interestingly, these six traits are not always the most comorbid or enriched ones with depression. For example, surprisingly, hypertension (enrichment *p*_*Hypertension*_ = 7.48 × 10^−833^) was not found genetically correlated with depression. In the resulting genetic network, we noticed that gastro-oesophageal reflux disease (GORD) had the highest number of connections with all other traits. More precisely, GORD was genetically correlated with eight other traits in the genetic network, thus forming a “hub” in our network (Figure 2).

**Table 3:** Pairwise significant genetic correlations. *Rg* denotes the genetic correlation coefficient; *SE* and *P* denote the standard error and test *p*-value for *Rg* respectively. The genetic correlation was computed using LDSC (14). See also Figure 2.

| Trait 1 | Trait 2 | $R_g$ | $SE$ | $P$ |
| --- | --- | --- | --- | --- |
| Anxiety | Depression | 0.8739 | 0.1678 | $1.90 \times 10^{-07}$ |
| Anxiety | Gastro-oesophageal Reflux | 0.4876 | 0.1190 | $4.17 \times 10^{-05}$ |
| Asthma | Gastro-oesophageal Reflux | 0.2047 | 0.0358 | $1.12 \times 10^{-08}$ |
| Asthma | Osteoarthritis | 0.2643 | 0.0668 | $7.65 \times 10^{-05}$ |
| Body Mass Index | Depression | 0.1140 | 0.0234 | $1.11 \times 10^{-06}$ |
| Body Mass Index | Gastro-oesophageal Reflux | 0.3007 | 0.0201 | $2.18 \times 10^{-50}$ |
| Body Mass Index | Inguinal Hernia | -0.2361 | 0.0302 | $5.35 \times 10^{-15}$ |
| Body Mass Index | Lipoprotein HDL | -0.2898 | 0.0300 | $4.71 \times 10^{-22}$ |
| Body Mass Index | Osteoarthritis | 0.4385 | 0.0437 | $1.17 \times 10^{-23}$ |
| Chronic Gastritis | Depression | 0.3735 | 0.0790 | $2.28 \times 10^{-06}$ |
| Chronic Gastritis | Gastro-oesophageal Reflux | 0.8434 | 0.0975 | $5.18 \times 10^{-18}$ |
| Depression | Gastro-oesophageal Reflux | 0.5221 | 0.0271 | $1.48 \times 10^{-82}$ |
| Depression | Osteoarthritis | 0.3556 | 0.0519 | $7.19 \times 10^{-12}$ |
| Depression | Suicidal Ideation Self Harm | 0.9687 | 0.1904 | $3.65 \times 10^{-07}$ |
| Gastro-oesophageal Reflux | Inflammatory Bowel Disease | -0.1065 | 0.0293 | $2.84 \times 10^{-04}$ |
| Gastro-oesophageal Reflux | Lipoprotein HDL | -0.1954 | 0.0246 | $1.72 \times 10^{-15}$ |
| Gastro-oesophageal Reflux | Osteoarthritis | 0.5348 | 0.0446 | $3.50 \times 10^{-33}$ |
| Hypertension | Lipoprotein HDL | -0.2728 | 0.0712 | $10^{-04}$ |

**Figure 2:**
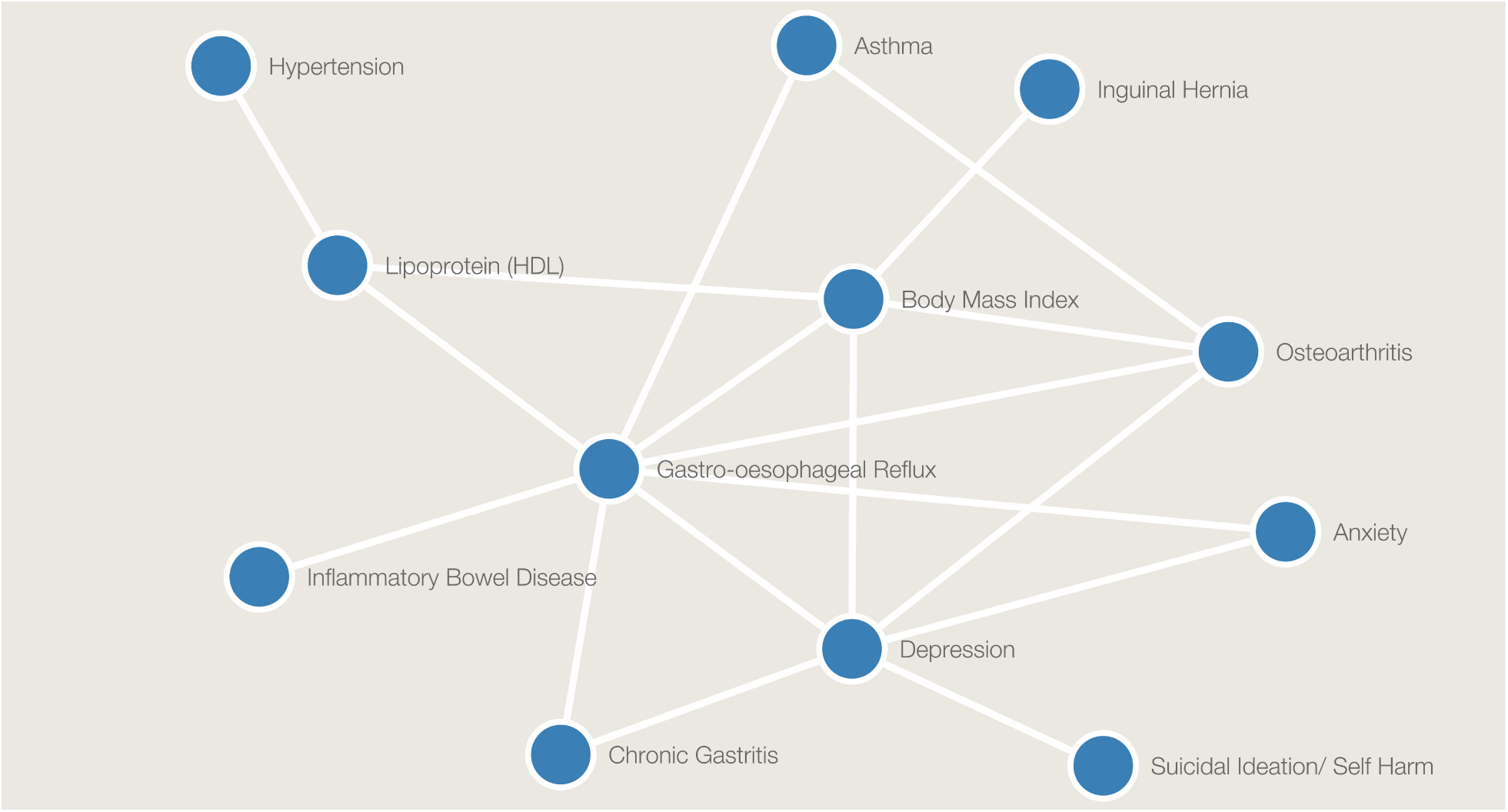
Genetic disease network of depression. For all connections observed in the phenotypic network of depression (figure 1), we evaluated the pairwise genetic correlation using LDSC(14). Edges in the graph show all significant genetic correlations between traits after correcting for multiple hypothesis testing.

### 3.3 Phenome-wide causal inference

For the 18 genetically correlated pairs of disorders discussed in the previous section, we used-Mendelian randomization as implemented in the bidirectional GSMR to test for the existence of putative causal association for each pair. Using a Bonferroni-corrected significance threshold equal to 1.39 × 10^−3^, we identified 18 traits as being potential causal exposures for an outcome, while using (nearly independent) associated SNPs as instrumental variables. Among the six traits genetically correlated with depression, we identified three causal risk factors for depression: GORD (169 independently associated SNPs with *b*_*xy*_ = 0.25 and *p* = 1.34 × 10^−30^); body mass index (BMI) (541 independently associated SNPs with *b*_*xy*_ = 0.17 and *p* = 4.08 × 10^−14^); and osteoarthritis (16 independently associated SNPs, *b*_*xy*_ = 0.10 and *p* = 7.98 × 10^−4^). Depression itself also appeared to have causal effects on both anxiety (*b*_*xy*_ = 0.62, *p* = 2.99 × 10^−09^, and *N*_*SNP*_ = 50) and GORD (*b*_*xy*_ = 0.11, *p* = 2.66 × 10^−11^, and *N*_*SNP*_ = 64). Recall that GORD was a hub in our genetic network and GORD is again a hub in the causality network (see Figure 3 and Table 4).

**Figure 3:**
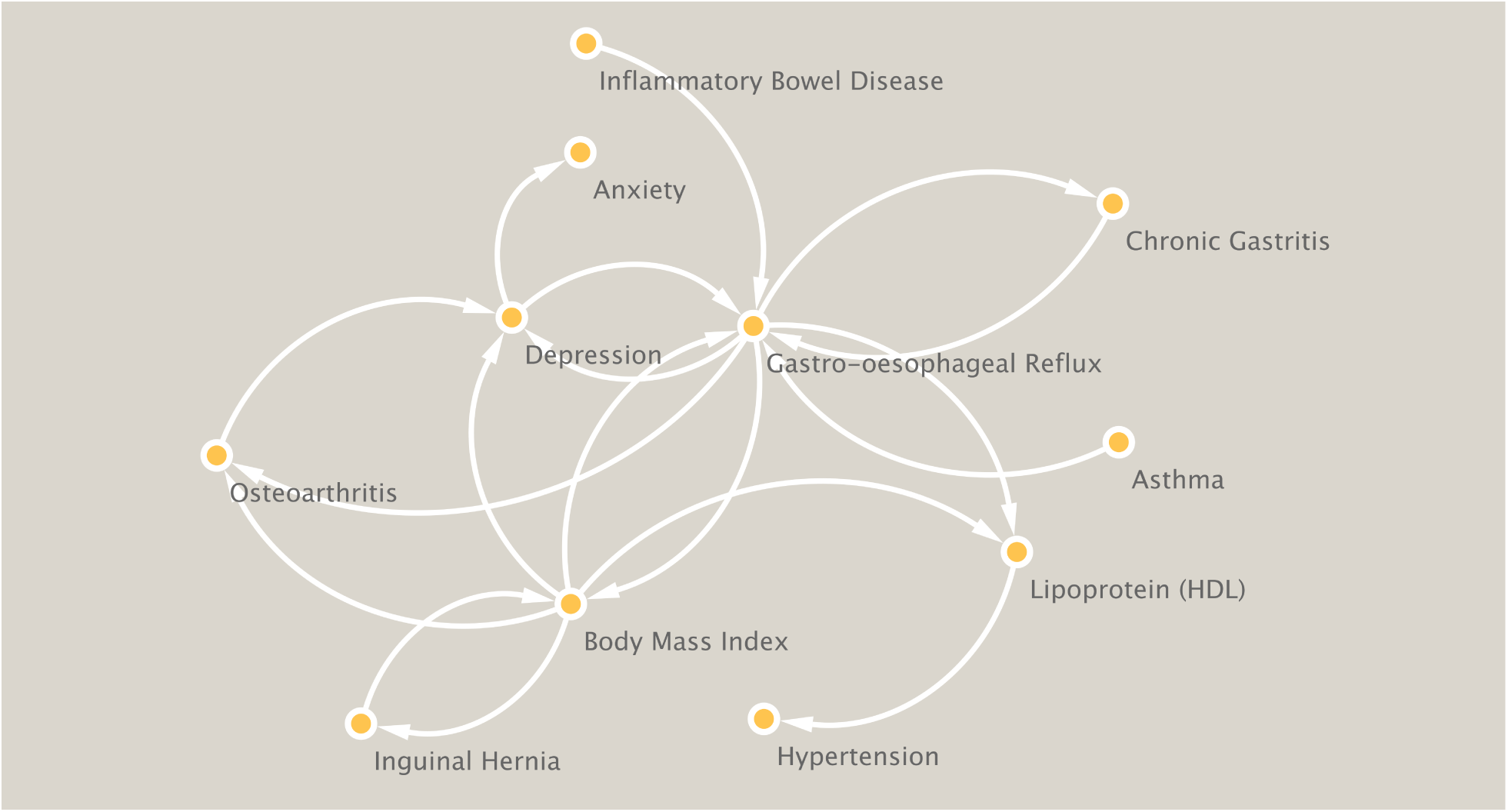
Causal relationships between traits that are comorbid with depression. We tested causal relationships between traits and depression using bidirectional Mendelian randomization for all genetically correlated pairs of disorders. Using SNP effects as instrumental variables, the resulting directed graph shows significant causalities between traits after correcting for multiple hypothesis testing.

**Table 4:**
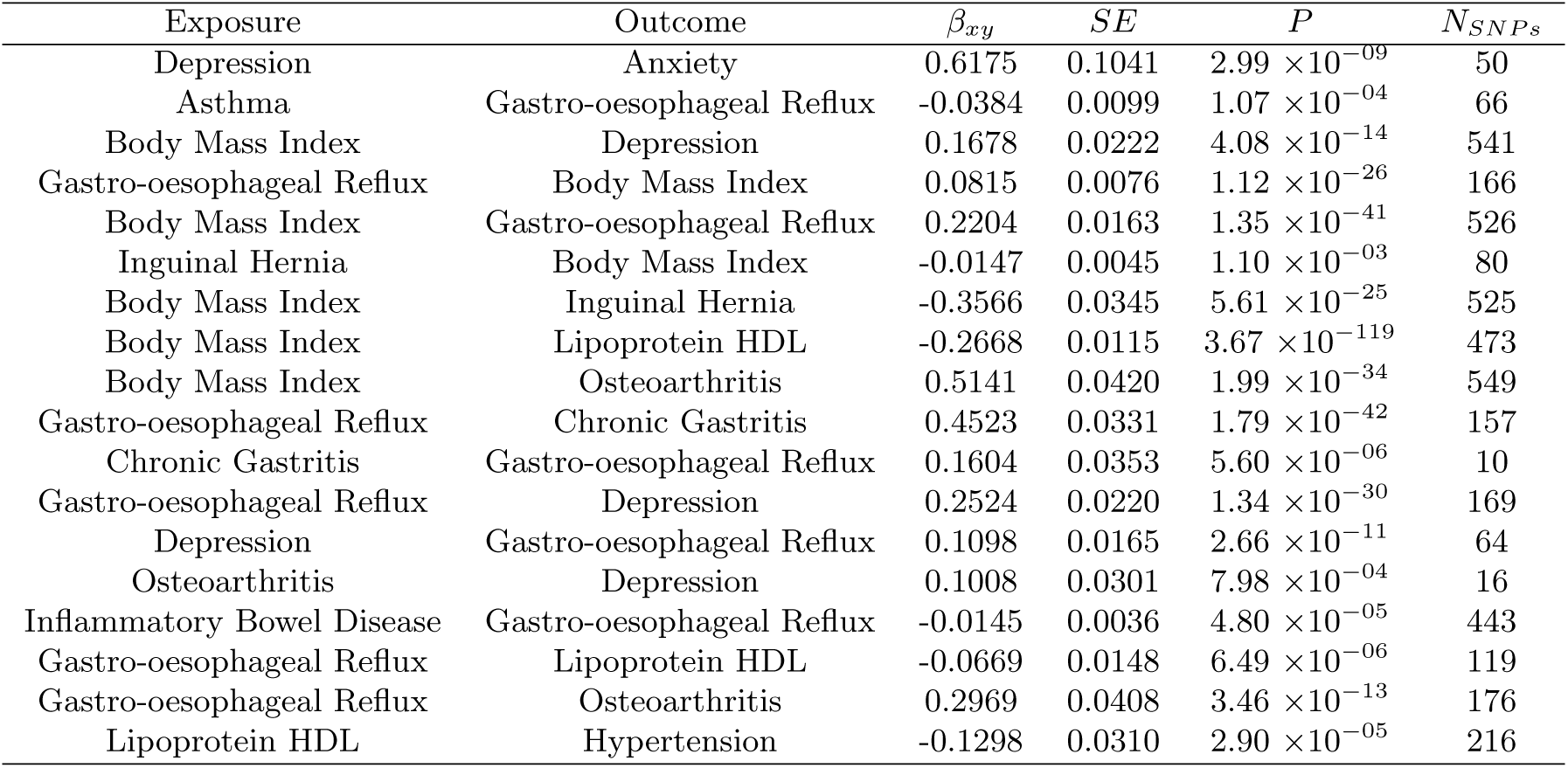
Causalities between exposures and outcomes from the Mendelian randomization analysis. *β*_*xy*_, *SE*, and *P* are effect size, corresponding standard error, and test *p*-value, respectively. *N*_*SNP s*_ is the number of independent SNPs used as instruments in the analyses. Mendelian randomization was carried out using the GSMR tool implemented in GCTA (111). See Figure 3 as well.

### 3.4 Pleiotropic variants underlying depression and potential causal exposures

For the three disorders that were found as potential immediate causal exposures of depression based on our Mendelian randomization analysis (GORD, BMI, and osteoarthrosis), we further looked for evidence of horizontal pleiotropy (see Table S3 for SNP-based pleiotropic results). We identified 522 SNPs showing genome-wide significant pleiotropic effects in either of the tested directions between depression and GORD. Only one of them also surpassed the Bonferroni-corrected threshold (*p* < 0.05/522 = 9.58 × 10^−5^) in the other direction and was therefore identified as pleiotropic. SNP rs11040813 (*p*_*pleio*1_ = 3.70 × 10^−05^, *p*_*pleio*2_ = 3.80 × 10^−08^) is located in region 11p15.4 close to gene *CNGA4*. For BMI, we identified a total of 10,667 SNPs showing genome-wide significant pleiotropic effects with depression in at least one of the tested directions. 103 of these SNPs were identified as pleiotropic (*p*_*pleio*1_ < 5.00 × 10^−08^ and *p*_*pleio*2_ < 4.69 × 10^−06^, or *p*_*pleio*2_ < 5.00 × 10^−08^ and *p*_*pleio*1_ < 4.69 × 10^−06^). The SNPs span regions 1p31.1, 10q25.1 and 16q12.2. 13 of the 103 SNPs showed genome-wide significant pleiotropic effects in both directions and all of them are located in region 1p31.1 (gene *LOC105378797*). For depression and osteoarthritis, 472 SNPs were picked up as genome-wide significant for one of the tested directions. However, none of them satisfied the necessary requirements to be a bidirectional pleiotropic SNP.

To seek higher power and better interpretability, we also carried out pleiotropic screening at a gene level by performing gene-based analyses in addition to SNP-based analyses (see Table 5 for gene-based pleiotropic analyses). For depression and GORD, a total of 18,172 protein coding genes were tested, making the genome-wide significance threshold less than 2.75 × 10^−06^. 58 genes were found significant at this threshold in at least one direction and eight were identified as pleiotropic (*p* < 0.05/58 = 8.62×10^−4^ in the other tested direction). Among these eight genes, the one that was most strongly associated with both directions was *TCF4* (*p*_*pleio*1_ = 4.18 × 10^−6^*andp*_*pleio*2_ = 1.67 × 10^−9^), followed by *HIST1H2BN* (*p*_*pleio*1_ = 7.34 × 10^−7^ and *p*_*pleio*2_ = 2.73 × 10^−6^). For the trait pair depression and BMI, we tested a total of 18,292 genes, resulting in a genome-wide gene significance threshold less than 2.73 × 10^−06^. Using this threshold, 391 genes showed pleiotropic effect in either direction. Five of them also showed a *p*-value smaller than 0.05/391 = 1.28 × 10^−4^ in the other direction and were identified as pleiotropic genes for the two traits. The list included *NEGR1* (*p*_*pleio*1_ = 1.29 × 10^−20^ and *p*_*pleio*2_ = 6.30 × 10^−6^) and *RAB27B* (*p*_*pleio*1_ = 7.44 × 10^−7^ and *p*_*pleio*2_ = c 9.47 × 10^−8^). Finally, for the trait pair depression and osteoarthritis, 18,230 protein coding genes were tested and 31 were found significant (*p* value below 2.74×10^−06^ in one of the tested directions). However, similar to the SNP-based analysis, none of the genes satisfied the necessary requirements to be classified as bidirectional pleiotropic genes.

**Table 5:** Pleiotropic genes between depression and its immediate risk factors. For all three immediate risk factors of depression, we looked into horizontal pleiotropic genetic variants shared between depression and the risk factor using IMRP (110). The analysis was done at both the SNP and the gene level. (See table S3 for SNP-based results.) GeneName, CHR, START, and STOP denote the gene and its genomic location; *N*_*SNP s*_ is number of SNPs analyzed for the respective gene; *Z* and *P* denote standardized pleiotropic effect sizes and test *p*-values; _*pleio*1_ stands for the SNP’s effect on depression, excluding the causality effect of the risk factor, and _*pleio*1_ stands for the SNP effect on the risk factor taking into account any causal effects from depression to the risk factor trait.

| GeneName | CHR | START | STOP | $N_{SNPs}$ | $Z_{pleio1}$ | $P_{pleio1}$ | $Z_{pleio2}$ | $P_{pleio2}$ |
| --- | --- | --- | --- | --- | --- | --- | --- | --- |
| Depression - GORD |  |  |  |  |  |  |  |  |
| *CCKBR* | 11 | 6280841 | 6293363 | 67 | 4.10 | $2.09 \times 10^{-05}$ | 5.18 | $1.14 \times 10^{-07}$ |
| *PGBD1* | 6 | 28249314 | 28270326 | 59 | 4.85 | $6.30 \times 10^{-07}$ | 4.06 | $2.41 \times 10^{-05}$ |
| *HIST1H2BN* | 6 | 27805544 | 27821533 | 32 | 4.82 | $7.34 \times 10^{-07}$ | 4.55 | $2.73 \times 10^{-06}$ |
| *ZSCAN26* | 6 | 28234788 | 28246001 | 18 | 4.87 | $5.56 \times 10^{-07}$ | 3.40 | $3.32 \times 10^{-04}$ |
| *TCF4* | 18 | 52889562 | 53303252 | 677 | 4.46 | $4.18 \times 10^{-06}$ | 5.91 | $1.67 \times 10^{-09}$ |
| *ZKSCAN4* | 6 | 28212404 | 28227030 | 33 | 4.99 | $2.94 \times 10^{-07}$ | 4.07 | $2.34 \times 10^{-05}$ |
| *BTN2A1* | 6 | 26458132 | 26476849 | 79 | 4.56 | $2.58 \times 10^{-06}$ | 4.67 | $1.54 \times 10^{-06}$ |
| *BTN3A2* | 6 | 26365387 | 26378548 | 97 | 5.03 | $2.49 \times 10^{-07}$ | 3.85 | $5.93 \times 10^{-05}$ |
| Depression - BMI |  |  |  |  |  |  |  |  |
| *IP6K1* | 3 | 49761728 | 49823973 | 137 | 5.73 | $5.07 \times 10^{-09}$ | 3.70 | $1.07 \times 10^{-04}$ |
| *RAB27B* | 18 | 52385097 | 52562747 | 681 | 4.81 | $7.44 \times 10^{-07}$ | 5.21 | $9.47 \times 10^{-08}$ |
| *DENND1A* | 9 | 126141933 | 126692423 | 1825 | 8.32 | $4.51 \times 10^{-17}$ | 3.70 | $1.08 \times 10^{-04}$ |
| *DNAJC11* | 1 | 6694228 | 6761966 | 201 | 4.74 | $1.07 \times 10^{-06}$ | 3.75 | $8.84 \times 10^{-05}$ |
| *NEGR1* | 1 | 71868625 | 72748533 | 2550 | 9.24 | $1.29 \times 10^{-20}$ | 4.37 | $6.30 \times 10^{-06}$ |

## 4 Discussion

Motivated by the highly heterogeneous nature of the phenotypic manifestations and genetic basis of depression and its many co-occuring disorders we embarked on a systematic evaluation of their phenotypic and genetic relationships seeking to gain insights into the biological mechanisms that may underlie comorbidities in depression. We uncovered intriguing phenotypic and genetic relationships between depression and GORD and BMI, as well as genes that could play a central role in governing the links between mental disorders and disorders that are not immediately thought of as relating to the nervous system.

Using ICD-10 records of the European samples in the UK Biobank, we began by constructing a (phenotypic) comorbidity network of depression. We mapped over six thousands ICD codes onto approximately four hundreds PheCodes of heritable traits. We discovered 12 traits that are often comorbid with depression, including mental disorders (for instance suicidal ideation and anxiety) we well as traits affecting the digestive system (abdominal hernia, gastritis and duodenitis, functional digestive disorders, diseases of esophagus), metabolic disorders (fluid and electrolyte disorders, lipid metabolism, obesity, etc.), asthma, hypertension, and osteoarthrosis. Many of these comorbidities have been previously observed in epidemiological studies, i.e., such as GORD (20), obesity (17), asthma (47), hypertension condition (34), and the more well-studied anxiety disorders (98) and suicidal ideation (29; 73).

Next, using GWAS summary statisitcs, we explored the genetic correlation between all highly comorbid pairs of disorders in the phenotypic network. This analysis resulted in a, somewhat less well-connected network (compared to the phenotype-based one), where we observed that traits that are phenotypically connected with depression do not necessarily share a common genetic basis with it. Interestingly, hypertension, one of the most enriched conditions among depression patients, did not show significant genetic correlation with depression. This implies that high comorbidity rates between depression and hypertension (or even other cardiovascular traits), may not be due to a shared heritable etiology. Instead, such phenotypic commorbidities may be mediated by other factors (18), such as drug effects of antidepressants (95; 45). This lack of genetic connections between depression and hypertension is concordant with the observation that these two traits show unique, although interactive, effects on cognitive functions and brain volumes as has been previously described (87; 67).

Significant genetic correlations have been validated by our results between depression and psychiatric traits (anxiety and suicidal ideation) (70; 27) We also identified links connecting depression with GORD, BMI, chronic gastritis, and osteoarthrosis, reinforcing the complexity of depression’s etiology. Strong clinical relationships between the digestive system and psychiatric conditions have been known for over 35 years (31; 100). Recent studies also provided further evidence supporting genetic commonalities between depression and various digestive traits (107), including GORD (79) and chronic gastritis (3). GORD was a “hub” in our genetic network and appears significantly correlated with BMI and osteoarthrosis, two traits that share a direct genetic basis with depression. GORD also creates connections between depression and hypertension, asthma, and IBD.

For all pairs of disorders that we found to be (significantly) genetically correlated we looked into potential causal relationships using Mendelian randomization. All the traits in the connected component of the genetic network (except suicidal ideation) remained connected in the causality network. We found three immediate risk factors for depression: GORD, BMI, and osteoarthrosis. Additionally, depression arose as a significant risk factor for anxiety and GORD. The other traits, although not directly linked to depression, appeared to be connected via intermediate nodes. GORD appeared as a hub and, along with BMI, could be acting to connect depression to all other digestive traits. Interestingly, the asthma node also pointed towards depression via GORD. Phenotypic comorbidity between asthma and GORD has been widely reported (12; 7), whereas, to the best of our knowledge, evidence for a causal link between these two disorders has not been suggested until now. We used SNP effects as instruments and we provided insights into possible relationships between asthma and GORD; biological explanations explaining such causal relationships might be due to the epithelial dysfunction in asthma, since such patients are more vulnerable to acid reflux (81). Our results also point towards a uni-directional causality from BMI to depression. This observation is supported by longitudinal studies, where obesity was found to increase the risk of depression (59). Many studies found that the distribution of depression incidence as a function of BMI is a U-shaped curve (61; 25; 109), indicating that not only obesity, but also any BMI abnormalities could result in a higher risk of depressive conditions. On the other hand, the relationship between depression and osteoarthrosis has not received as much attention as the other trait pairs, perhaps due to the fact that the latter is strongly age-dependent; a relatively recent PheWAS discovered an association between the genetic risk score for depression and osteoarthrosis in the UK biobank samples (72). On the clinical side, links between osteoarthrosis/arthrosis and depressive traits are more frequently explained by the pain caused by such conditions (58; 104; 94), which is also in concordance with our finding that osteoarthrosis indicated a causal effect on depression, but not the other way around.

In order to further explore the biological mechanisms underlying connections between the afore-mentioned traits and depression, we investigated genetic horizontal pleiotropic effects at the SNP and gene levels for depression and its three most closely correlated traits: GORD, BMI, and osteoarthrosis. We identified several multi-functional genetic regions and genes that play a role in the etiology of depression and its comorbidities. Our gene-based pleiotropy analysis of depression and GORD highlighted the role of transcription factor 4 (*TCF4*), which regulates the differentiation of various cell types and underlies the etiology of both diseases. This gene has also been reported to have an impact in depression and insomnia (15). Beyond depression and GORD, associations between *TCF4* and many other mental and physical conditions have been widely studied; such conditions include cognitive ability, autism, the development of certain cancer types, as well as rare diseases such as Pitt-Hopkins syndrome (33; 74; 51; 56; 5). Moreover, *TCF4* has been identified as one of the strongest genetic biomarkers for schizophrenia susceptibility in multiple genetic studies (44; 90; 52). Animal experiments suggest that the over-expression of this gene in the brain results in functional impairments that constitute schizophrenic symptoms (13) and that such effects further interact with different exposure conditions (8). Interestingly, in patients with depression, a decreased *TCF4* expression at the mRNA and protein level was observed when compared to healthy controls (71). Our results indicate that, beyond schizophrenia, it may also be worthwhile to further investigate the pathology of *TCF4* in depressive conditions.

One of the top genes that was uncovered from the pleiotropic analysis between depression and BMI was *NEGR1* (Neuronal Growth Regulator 1). This is a known obesity-associated gene, whose effect has been verified in multiple populations (66; 76; 83; 85). *NEGR1* regulates neuronal control of food intake (9; 53) and along with *FTO*, which was also picked up in the SNP-based pleiotropic analysis, have been reported as having an impact on eating disorders (68; 35). Given its high expression level in the brain, its connections with psychiatric traits has been recently investigated and *NEGR1* has been associated with a wide range of mental traits. Apart from eating related disorders, it also plays a role in depression (43), cognitive performance (54), and Alzheimer’s disease (77). As a functional validation, increased depressive, anxiety- and autistic-like behaviors were observed in *NEGR1*-deficient mice, along with reduced hippocampal neurogenesis and disruption in cortical development (78; 92). Finally, as a putative tumor suppressor, *NEGR1* has also been identified to be a down-regulated gene in various cancer cells (50; 93), including ovarian cancer (86; 21) and breast cancer (10; 82).

Regarding limitations for our study, first, our results are based on analysis of the UK Biobank and sample inclusion in the UK biobank is volunteer-based so might not represent the true underlying population distribution. However, most of the connections observed in our work have also been previously reported in epidemiological studies which provides further support for our findings. Second, our results used GWAS summary statistics, which exhibit some variance in their respective power. Additionally, while most of the GWAS that we used were performed on European populations, the GWAS for electrolyte disorders (GWAS of sodium and potassium Levels) was based on the Qatar population.

## 5 Conclusion

In this paper we systematically evaluated phenotypic and genetic connections between depression and other disorders and looked into their causality relationships. We also sought to uncover genes with potentially pleiotropic effects across depression and highly correlated disorders. Our results suggest that beyond phenotypic comorbidities, depression shows genetic connections with multiple psychiatric and non-psychiatric traits and we identify multiple genetic variants that could be linking mental health disorders to disorders of other systems. Such common genetic background may help explain the heterogeneity of the symptoms of depressive patients and help further sub-typing of depression. Our work highlights the value of trans-diagnostic approaches towards uncovering insights into the the etiology of psychiatric disease.

## Data Availability

All data produced in the present study are available upon reasonable request to the authors

## 6 Supplementary table legends

**Table S1. Parent phecodes and number of cases in our analysis**. After pre-processing, we mapped 6,300 ICD-10 codes onto 412 parent phenodes, including depression. The table is a detailed list of final phecodes used in the analyses and the number of cases after quality control.

**Table S2. Edges in the phenotypic network**. We computed pairwise phenotypic correlation between all phecodes included in our study. We extracted the top 1% highly-correlated pairs. We further filtered out all immediate neighbour nodes of depression and the network spanning these nodes. This network includes a total of 56 edges (listed in the table). Edge weights are the phenotypic correlations between two nodes, defined as the cosine of the angle formed by the two phenotype vectors.

**Table S3. SNP-based pleiotropic results for depression and GORD, and depression and BMI**. Pleiotropic SNPs identified between depression and its immediate risk factors (GORD and BMI) were identified by the Mendelian randomization analysis.

